# Cardiac sympathetic degeneration informs the duration of the prodromal stage of body-first Lewy body disease

**DOI:** 10.64898/2026.01.28.26344978

**Authors:** Casper Skjærbæk, Ole Lajord Munk, Katrine B. Andersen, Anushree Krishnamurthy, Karoline Knudsen, Thea Lillethorup, Sang-Won Yoo, Dong-Woo Ryu, Yoonsang Oh, Seunggyun Ha, Joong-Seok Kim, Per Borghammer, Jacob Horsager

## Abstract

Body-first Lewy body disease (LBD) is hypothesized to begin in the peripheral autonomic nervous system, years before nigrostriatal involvement. Isolated REM sleep behaviour disorder (iRBD) is considered prodromal body-first LBD, but the duration of the prodromal phase remains unknown. We aimed to determine the progression rate of cardiac sympathetic denervation using [^123^I]meta-iodobenzylguanidine (MIBG) scintigraphy and employ the resulting curves to estimate the prodromal period of body-first LBD.

We analysed longitudinal MIBG and dopaminergic imaging data from three cohorts: early Parkinson’s disease (PD) patients from KPD, Korea (KPD-PD; n=195); de novo PD (n=74) and iRBD (n=54) patients from the PACE cohort, Aarhus, Denmark; and DaT SPECT data from PPMI PD (n=426) and iRBD (n=37). Heart-to-mediastinum ratios (MIBG) and putamen-to-occipital ratios (DaT SPECT) were converted to percent of normal value (healthy control mean). Patients were binned into quartiles according to baseline imaging data and progression curves were constructed by using median decline rates within each quartile. Onset of cardiac sympathetic (peripheral) and nigrostriatal dopaminergic (central) neurodegeneration were determined for iRBD patients by back-extrapolation from baseline imaging values.

Longitudinal MIBG trajectories were highly consistent across the KPD and PACE cohorts, showing a rapid early decline (∼10%-points/year) followed by gradual slowing, reaching 50% of normal value after ∼5 years and 25% after ∼10 years. PACE-iRBD patients displayed severe baseline cardiac sympathetic loss (median 21.3% of normal), corresponding to an estimated onset of peripheral neurodegeneration 11.3 years prior to study enrolment. Dopaminergic decline was slightly slower, reaching 50% of normal value after ∼8 years and 25% after ∼15 years. PACE-iRBD patients exhibited mild baseline dopaminergic deficit (median 81.6% of normal), indicating onset of nigrostriatal degeneration 2.7 years prior to enrolment. Thus, cardiac sympathetic degeneration preceded nigrostriatal involvement by 8.6 years. Based on baseline dopaminergic degeneration in PACE-PD patients, predicted time to phenoconversion for PACE-iRBD patients was 8.4 years. The combined model estimated the total prodromal period in body-first LBD to exceed 19 years.

In conclusion, our study suggests that cardiac sympathetic degeneration begins more than a decade before study enrolment in iRBD subjects and nearly two decades before LBD diagnosis. Therefore, MIBG scintigraphy is a robust biomarker for detecting the earliest measurable neurodegeneration of body-first LBD and may be integrated in biological staging of α-synucleinopathies. Furthermore, our findings have implications for drug trial design in SAA-positive individuals, and for identifying patients at the optimal window for disease-modifying therapies.

## Introduction

Lewy body disease (LBD) comprises the clinical phenotypes Parkinson’s disease (PD) and Dementia with Lewy bodies (DLB), characterized by intraneuronal accumulation of α-synuclein. These inclusions are also found in postmortem examinations of patients with isolated REM-sleep behaviour disorder (iRBD)^1^, a parasomnia frequently encountered in LBD. Nearly all iRBD patients develop PD or DLB within 15 years^2^. Therefore, iRBD is considered a prodromal manifestation of LBD and the strongest prodromal marker of incipient PD^3^.

LBD patients exhibit neuronal degeneration of multiple neurotransmitter systems in the brain and the peripheral autonomic nervous system. Cardiac sympathetic neurons are among the most vulnerable, but a striking heterogeneity in the timing of this degeneration is seen. At diagnosis, some LBD patients already show severe cardiac sympathetic denervation, whereas others are completely normal - despite overt brain involvement. Among other observations, this has prompted the idea that LBD comprises two main subtypes^4,5^: a *brain-first subtype* initially affecting the brain and later the peripheral autonomic nervous system, and a *body-first subtype* initially affecting the peripheral autonomic nervous system years before the brain. Cardiac denervation as a marker of body-first LBD is further supported by a recent autopsy study demonstrating that brainstem-predominant Lewy body profiles (i.e. body-first) were associated with severe cardiac sympathetic denervation, whereas amygdala-predominant profiles (i.e. brain-first) showed much more preserved innervation^6^. This finding strongly supports that iRBD patients are mainly prodromal body-first LBD patients with brainstem-predominant Lewy pathology, since 90-95% of iRBD cases display severe cardiac sympathetic denervation on imaging^7-9^.

[^123^I]meta-iodobenzylguanidine (MIBG) scintigraphy is a robust tool to evaluate cardiac sympathetic innervation and has been validated against postmortem myocardial tyrosine hydroxylase immunostaining^10,11^. Late static images reflect the uptake and storage of MIBG, i.e., a sympathetic functional entity^12^. MIBG scintigraphy can distinguish PD from atypical parkinsonism and serve as a supportive/indicative biomarker in the diagnostic criteria for PD and DLB, respectively ^13,14^.

Nevertheless, while an extensive literature on dopaminergic degeneration exists, the progressive peripheral autonomic degeneration has received limited attention. Therefore, a temporal understanding of the onset and subsequent progressive loss of autonomic neurons is needed. Here, we examine the progressive cardiac denervation using MIBG scintigraphy in large cohorts of iRBD and PD patients. We aimed to model the time course of cardiac sympathetic denervation and estimate how it relates to the onset of nigrostriatal neurodegeneration. This strategy enables an accurate estimate of the prodromal period in body-first LBD.

## Methods

### Participants

Subject characteristics are presented in **Table 1**. We included data from three separate cohorts:

i. A Korean Nationwide Parkinson’s Disease cohort^15^, recruited at movement disorder clinics of KPD St. Mary’s Hospital and Yeouido St. Mary’s Hospital (designated KPD), comprising early PD patients with longitudinal MIBG (KPD-PD).
ii. A Lundbeck Foundation Parkinson’s Disease Research Center cohort, recruited at Aarhus University Hospital, Denmark (designated PACE), comprising de novo PD patients and iRBD patients (PACE-PD and PACE-iRBD) with longitudinal MIBG and baseline dopaminergic imaging ([^18^F]-FDOPA PET or [^18^F]-FE-PE2I PET).
iii. Data extracted from the Parkinson’s Progression Marker Initiative cohort (designated PPMI), comprising early PD patients (PPMI-PD) and iRBD patients (PPMI-iRBD) with longitudinal dopamine transporter (DaT) imaging. Data for this paper were acquired from the PPMI database on August 18^th^, 2025. To estimate dopamine transporter loss over time, we included patients from the PPMI dataset with baseline DaTscans obtained between 2010-07-01 and 31-12-2019.

**Table 1.** Demographics and clinical characteristics of the patient cohorts.

|  | **PACE** |  | **KPD** | **PPMI** |  |
| --- | --- | --- | --- | --- | --- |
|  | **PD** | **iRBD** | **PD** | **PD** | **iRBD** |
| **n** | 74 | 54 | 195 | 426 | 37 |
| **Sex (F/M)** | 20/54 | 8/46 | 97/98 | 154/272 | 6/31 |
| **Age at baseline (years)** | 67.0 (8.2) | 66.8 (7.6) | 67.7 (8.9) | 61.7 (9.7) | 69.1 (5.3) |
| **Time since diagnosis at baseline (years)** | 0.59 (0.82) | 1.00 (1.13) | 0 | 0.80 (1.20) |  |
| **Symptom duration at baseline (years)** | 2.13 (1.59) | 6.43 (4.77) | 1.16 (1.08) | 2.18 (2.18) |  |
| **UPDRS-III at baseline** | 19.01 (10.20) | 5.64 (6.87) | 19.06 (9.32) | 21.79 (9.43) | 4.38 (3.82) |
| **HoeHN & Yahr stage at baseline (0/1/2/3/4)** | 0/28/40/5/0 |  | 0/61/120/14/0 | 0/179/245/2/0 |  |
| **Baseline imaging (n)** | MIBG =73<br>FDOPA = 37<br>PE2I = 33 | MIBG = 54<br>FDOPA = 21<br>PE2I = 32 | MIBG =195 | DaT SPECT = 426 | DaT SPECT = 37 |
| **Follow-up imaging (n)** | MIBG = 44<br>FDOPA = 22<br>PE2I = 22 | MIBG = 21<br>FDOPA = 13<br>PE2I = 9 | MIBG = 195 | DaT SPECT = 426 | DaT SPECT = 37 |
| **Follow-up time (baseline to FU, years)** | 3.12 (0.37) | 3.32 (1.18) | 2.27 (0.53) | 3.45 (1.05) | 3.63 (1.05) |
| **MIBG at baseline (H/M late, % of normal)** | 34.4%<br>[17.9, 67.7] | 21.3%<br>[14.0, 38.9] | 33.7%<br>[19.2, 55.2] |  |  |
| **MIBG at follow-up (H/M late, % of normal)** | 26.0%<br>[18.2, 49.2] | 16.7%<br>[7.59, 32.1] | 24.6%<br>[15.2, 50.5] |  |  |
| **Dopamine imaging at baseline (lowest putamen, % of normal)** | 36.7%<br>[29.2, 44.8] | 81.6%<br>[70.6, 94.0] |  | 31.4%<br>[24.1, 39.3] | 54.8%<br>[45.9, 67.3] |
| **Dopamine imaging at follow-up (lowest putamen, % of normal)** | 25.0%<br>[17.4, 32.5] | 69.5%<br>[60.1, 82.5] |  | 26.1%<br>[19.3, 32.3] | 50.9%<br>[36.8, 72.8] |

*Data presented as mean(SD) or median[IQR]. Symptom duration is defined as the time elapsed since self-reported onset of motor symptoms in PD or onset of RBD symptoms in iRBD.*

Each study was approved by the relevant local research ethics committees, and all participants provided written informed consent. PD was diagnosed according to the Movement Disorder Society (MDS) diagnostic criteria in the KPD and PACE cohorts^14^. iRBD was diagnosed according to ICSD-3 criteria, based on video-polysomnography as specified in the AASM manual^16^. Patients with cardiac diseases with suspected impact on MIBG uptake, including larger myocardial infarctions and diabetic autonomic neuropathy, were excluded. In the PACE-iRBD dataset, patients diagnosed with DLB or PD within 6 months of the baseline scans were excluded from all analyses (1 DLB, 1 PD). From the PPMI dataset, PD patients with LRRK2 and SNCA mutations were excluded due to their overrepresentation in the dataset relative to the expected background prevalence.

### Imaging protocols

#### KPD

MIBG imaging was performed with a dual-head camera equipped with a low-energy, high-resolution collimator (Siemens, Germany). A late static image was obtained 2 hours after injection of 111 MBq.

#### PACE

MIBG images were obtained 3.5 hours after injection of 111 MBq on a dual-head camera equipped with a low-energy, high-resolution collimator (Siemens Symbia SPECT/CT, Germany). [^18^F]-FDOPA PET protocol have been described in detail previously^5^. In brief, carbidopa was administered 1 hour prior to injection. Following a 6 min transmission scan, PET data were acquired on a ECAT high-resolution research tomograph (Siemens/CTI) from 70-90 minutes after injection of 110MBq [^18^F]-FDOPA. [^18^F]-FE-PE2I PET was obtained on a Siemens Biograph Vision 600 PET/CT (Siemens Healthcare, Erlangen, Germany) from 30-40 min after injection of 100 MBq tracer. Dopaminergic imaging data were normalized to MNI space with rigid matching of the subject’s PET to a T1 anatomical MRI performed on either a 3T Siemens SKYRA magnetic resonance system or GE healthcare SIGNA PET/MR system.

#### PPMI

The dopamine transporter imaging ([^99m^Tc]-TRODAT-1 or [^123^I]Ioflupane) protocol can be retrieved here: https://www.ppmi-info.org/study-design/research-documents-and-sops. Healthy controls with visually abnormal DaT-scans or known genetic mutation were excluded. PD patients with visually normal DaT-scans were excluded. For subjects with multiple available DaT-scans, only the first and last available scans were used for analyses.

### Data analyses

#### Dopaminergic imaging

[^18^F]-FDOPA and [^18^F]-FE-PE2I PET data from PACE were analysed in PMOD. Volumes-of-interest were defined in the putamen and occipital cortex using Hammers N30R83 atlas. Similarly, putamen and occipital cortex data were extracted from the PPMI database. Specific putamen-to-occipital ratios were calculated as:

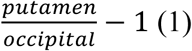

It has previously been shown that the putamen-to-occipital ratios show good correlation to the FDOPA influx constant (K_i_) obtained through full tracer kinetic modelling^17^. Putamen-to-occipital ratios were age-corrected with a linear model based on in-house reference material of healthy controls. The most affected putamen was used for analyses. Only PD patients with visually abnormal putamen on dopamine imaging were included.

#### MIBG scintigraphy

Regions-of-interest of the heart and mediastinum were manually defined, and average pixel counts were extracted to calculate heart-to-mediastinum (H/M) ratios. No age-correction was applied, as MIBG H/M ratios did not correlate with age in healthy control subjects.

#### Conversion to percentage of normal value

Specific putamen-to-occipital ratios were converted for [^18^F]-FDOPA, [^18^F]-FE-PE2I, and DaT SPECT, separately as:

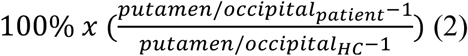

Dopaminergic tracers were age-adjusted and normalized to percentage-scale separately before combining the normalized data to a common percentage-scale.

MIBG H/M ratios were converted to percentage of healthy control mean as:

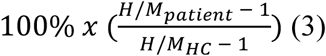

In the following, all percentages denote percentage of normal value (healthy control mean), i.e., *not* percentage loss.

To study the longitudinal dopaminergic and cardiac sympathetic degeneration, data was binned to quartiles based on baseline imaging result as follows: Q1>75%, Q2=50-75%, Q3=25-50%, and Q4<25% of normal value. Annual decline rates were calculated as:

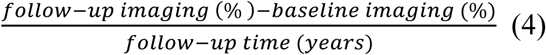

Importantly, this decline rate expresses %-points lost per year relative to the normal value, and not the percental specific-binding-ratio loss as reported in most other studies. For PPMI data, baseline and last visit data were used for analyses. Baseline results and decline rates for each quartile are shown for dopaminergic imaging and MIBG in **Table 2**. We excluded patients with increasing imaging values at follow-up for three reasons. First, to exclude patients who have not entered the neurodegenerative phase (for the respective imaging modality). Second, to avoid noise in the lower range of the curve, where progression approaches zero due to a floor effect. Third, to get a conservative estimate of the prodromal LBD period and hence underestimating, rather than overestimating, the true prodromal period. See the discussion section for more details.

**Table 2.**
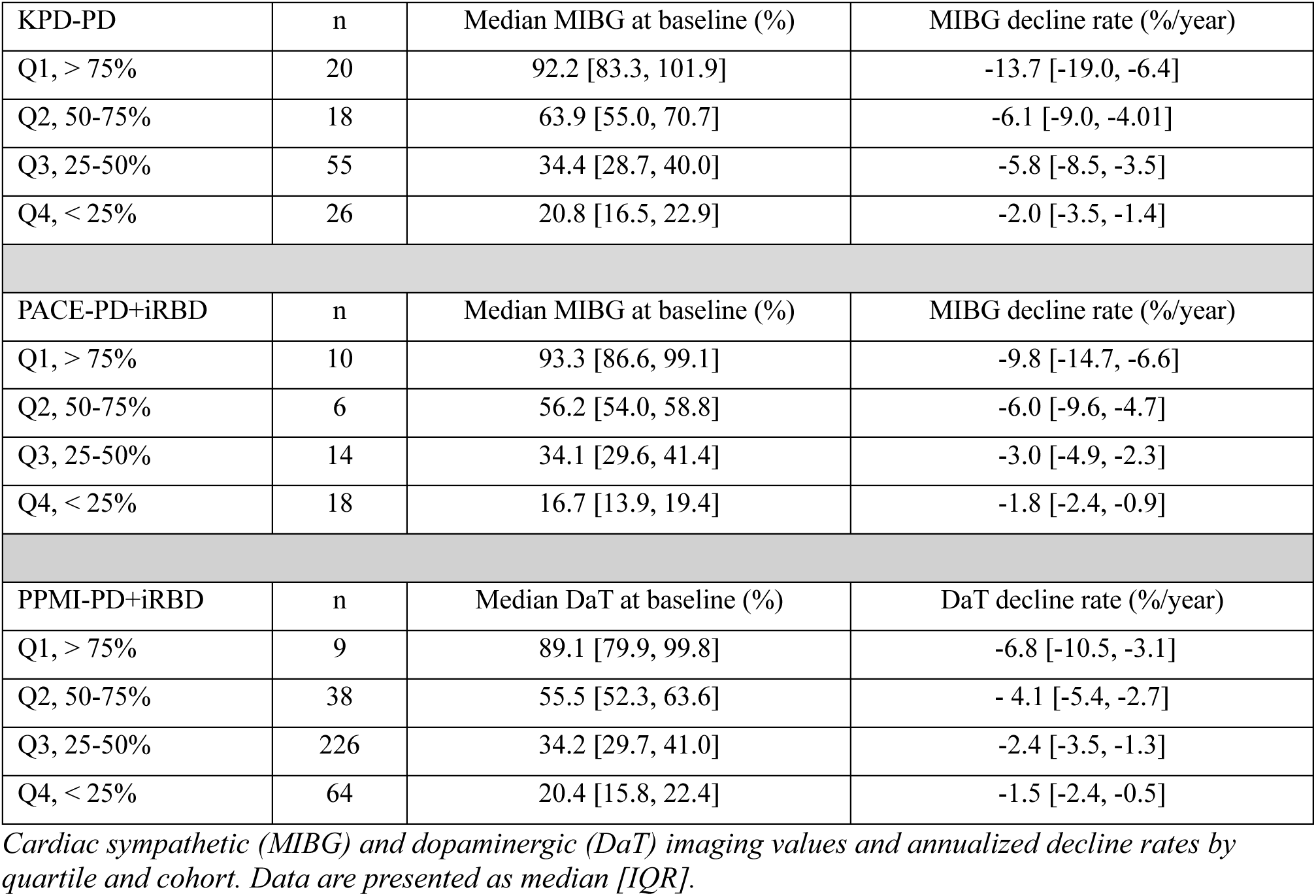
Baseline MIBG and dopamine imaging values and annualized decline rates.

| KPD-PD | n | Median MIBG at baseline (%) | MIBG decline rate (%/year) |
| --- | --- | --- | --- |
| Q1, > 75% | 20 | 92.2 [83.3, 101.9] | -13.7 [-19.0, -6.4] |
| Q2, 50-75% | 18 | 63.9 [55.0, 70.7] | -6.1 [-9.0, -4.01] |
| Q3, 25-50% | 55 | 34.4 [28.7, 40.0] | -5.8 [-8.5, -3.5] |
| Q4, < 25% | 26 | 20.8 [16.5, 22.9] | -2.0 [-3.5, -1.4] |
| PACE-PD+iRBD | n | Median MIBG at baseline (%) | MIBG decline rate (%/year) |
| Q1, > 75% | 10 | 93.3 [86.6, 99.1] | -9.8 [-14.7, -6.6] |
| Q2, 50-75% | 6 | 56.2 [54.0, 58.8] | -6.0 [-9.6, -4.7] |
| Q3, 25-50% | 14 | 34.1 [29.6, 41.4] | -3.0 [-4.9, -2.3] |
| Q4, < 25% | 18 | 16.7 [13.9, 19.4] | -1.8 [-2.4, -0.9] |
| PPMI-PD+iRBD | n | Median DaT at baseline (%) | DaT decline rate (%/year) |
| Q1, > 75% | 9 | 89.1 [79.9, 99.8] | -6.8 [-10.5, -3.1] |
| Q2, 50-75% | 38 | 55.5 [52.3, 63.6] | - 4.1 [-5.4, -2.7] |
| Q3, 25-50% | 226 | 34.2 [29.7, 41.0] | -2.4 [-3.5, -1.3] |
| Q4, < 25% | 64 | 20.4 [15.8, 22.4] | -1.5 [-2.4, -0.5] |

*Cardiac sympathetic (MIBG) and dopaminergic (DaT) imaging values and annualized decline rates by quartile and cohort. Data are presented as median [IQR].*

The median decline rate from Q1 subjects determined the upper part of the slope, i.e. from MIBG = 100% and Time = 0 until the median imaging value of Q2 was reached on the MIBG-axis (y-axis). The median decline rate for Q2 then determined the subsequent part of the slope until the median imaging value of Q3 was reached on the MIBG-axis and so forth. The curve was extrapolated to reach MIBG = 0%. More information about this method and individual data points are shown in **Supplementary Figures 1A-C and 2A-C.**

### Statistics

Descriptive statistics and graphical visualisation of imaging data were performed in *R* (version 4.5.1; *R* Core Team, 2023) under macOS (version 26.0.1) using the *tidyverse* framework for data visualisation.

## Results

### Patient cohort characteristics

We collected longitudinal MIBG data from a total of 260 LBD patients from three cohorts: the KPD-PD cohort (n = 195), PACE-PD cohort (n = 44) and PACE-iRBD cohort (n = 21). For comparison, we assembled longitudinal dopaminergic imaging data from the PPMI-PD (n = 426) and PPMI-iRBD cohort (n = 37).

Normative MIBG data for the KPD cohort were acquired from 25 healthy control subjects, mean age 67.4 ± 4.8 years, 76% females. For the PACE cohort, normative MIBG data were acquired from 42 healthy control subjects, mean age 66.0 ± 10.6 years, 48% females. Normative dopaminergic imaging data in the PACE cohort were acquired for each modality: [^18^F]-FDOPA PET from 20 healthy subjects, mean age 63.5 ± 6.4 years, 20% females; [^18^F]-FE-PE2I PET from 54 healthy control subjects, mean age 66.2 ± 10.3 years, 52% females. Part of the longitudinal KPD-PD data, baseline PACE data, and part of the normative imaging data have been published previously^5,7,18^. Normative dopaminergic imaging data from the PPMI cohort included 184 healthy controls, mean age 60.4 ± 11.4 years, 37% females.

### Time course of cardiac sympathetic denervation (MIBG)

The median cardiac sympathetic deficit at baseline measured with MIBG was very similar in the KPD-PD (33.7% of normal value) and PACE-PD (34.4%) patients, and lower in the PACE iRBD patients (21.3%) (**Table 1**). Cardiac sympathetic denervation progressed very similarly in the KPD and PACE cohorts (**Fig. 1**). Therefore, the curves were averaged. The average curve exhibited the fastest decline in the upper part, reaching approximately 50% of normal value after 5 years, followed by a gradually slower decline rate reaching 25% after 10 years, and 0% after 20-25 years. This translates into a median annual decline rate of ∼10% points in PD patients with baseline MIBG within the normal range (**Table 2**). With a median MIBG of 21.3% of normal value in PACE-iRBD patients, the resulting period of cardiac sympathetic degeneration can therefore be estimated to antedate baseline enrolment by at least 11.3 years (**Fig. 2**).

**Figure 1.**
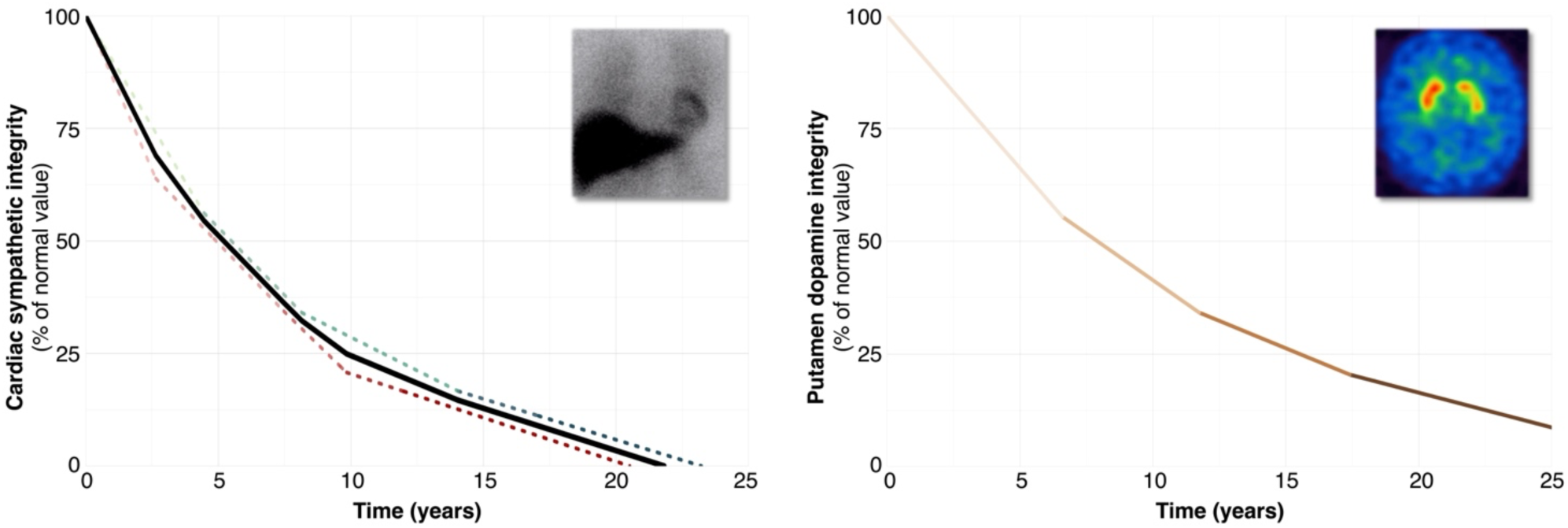
*Left:* Longitudinal decline of cardiac sympathetic integrity (assessed by MIBG) in two independent cohorts (KPD = red, PACE = green/blue, MEAN = black). Changing colour gradient along the curves indicates transitions between successive quartiles. *Right:* Longitudinal decline of putamen dopamine integrity, assessed by PPMI DaT imaging data (combined PD and prodromal). Notice the similarity between the two curves, although the MIBG curve seems to decline slightly faster. The MIBG curve reach 50% after 5 years and 25% after 10 years, whereas the DaT curve reach 50% of normal value after 8 years and 25% after 15 years.

**Figure 2.**
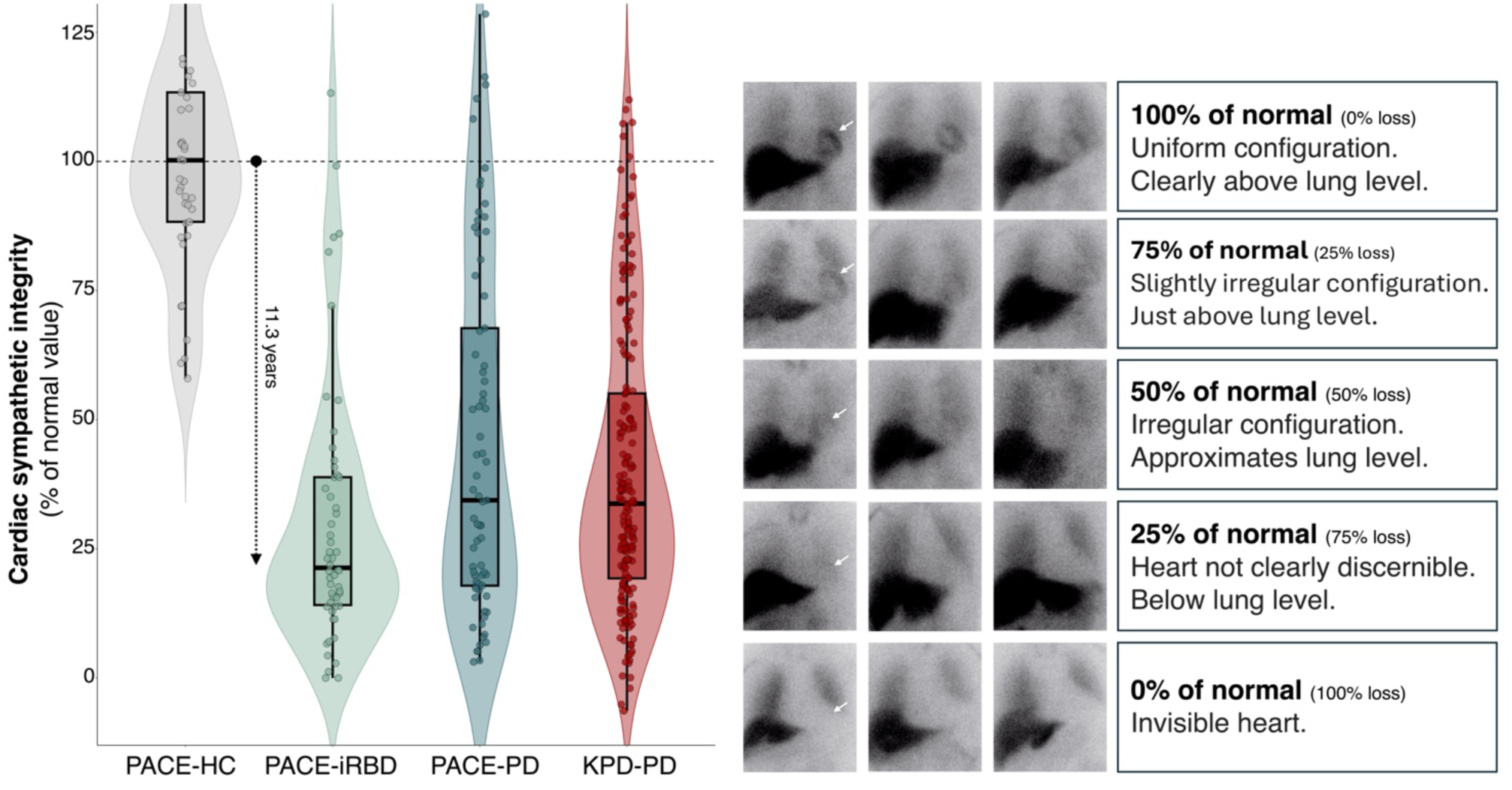
*Left:* Baseline cardiac sympathetic integrity (MIBG late H/M) data presented as % of normal value. The two PD cohorts show similar cardiac sympathetic degeneration. The PACE-iRBD cohort shows severe cardiac degeneration, median = 21.3%, reflecting a preceding degenerative phase of 11.3 years. Five PACE-HCs with values high in the normal range are not presented on the figure. *Right*: Three representative MIBG images are shown from each degenerative stage - from normal (100%) to complete denervation (0%). A guideline for qualitative interpretation of MIBG images is provided. White arrows signify location of the heart.

### Time course of nigrostriatal dopaminergic denervation

Median dopaminergic deficits were similar in the PPMI-PD (31.4%) and PACE-PD (36.7%) cohorts, whereas the iRBD patients showed more pronounced dopamine deficit in the PPMI-iRBD subjects (54.8%) compared to PACE-iRBD subjects (81.6%).

The time course of dopaminergic degeneration, measured from PPMI-PD and PPMI-iRBD data, was slightly slower that seen for MIBG. The curve showed an initial rapid decline, and then slower progression in the lower ranges, approximating 50% of normal value after 8 years, 25% after 15 years, and 0% after 25+ years (**Fig. 1**). With a median dopaminergic imaging level of 81.6% of normal value in PACE-iRBD patients, the average onset of dopaminergic degeneration can therefore be estimated to antedate baseline imaging by 2.7 years (**Fig. 3**). The time gap between onset of cardiac sympathetic degeneration and nigrostriatal dopaminergic degeneration was therefore 8.6 years (**Fig. 4**). Mean duration of RBD-symptoms in the PACE-iRBD cohort was 6.4 years, suggesting that RBD started *after* cardiac sympathetic degeneration but *before* nigrostriatal degeneration. The more pronounced baseline dopaminergic deficit in PPMI-iRBD subjects, resulted in an estimated onset of dopaminergic degeneration 6.8 years prior to baseline imaging. As the PPMI does not include MIBG, the time gap between onset of dopaminergic and cardiac sympathetic degeneration could not be determined for this cohort.

**Figure 3.**
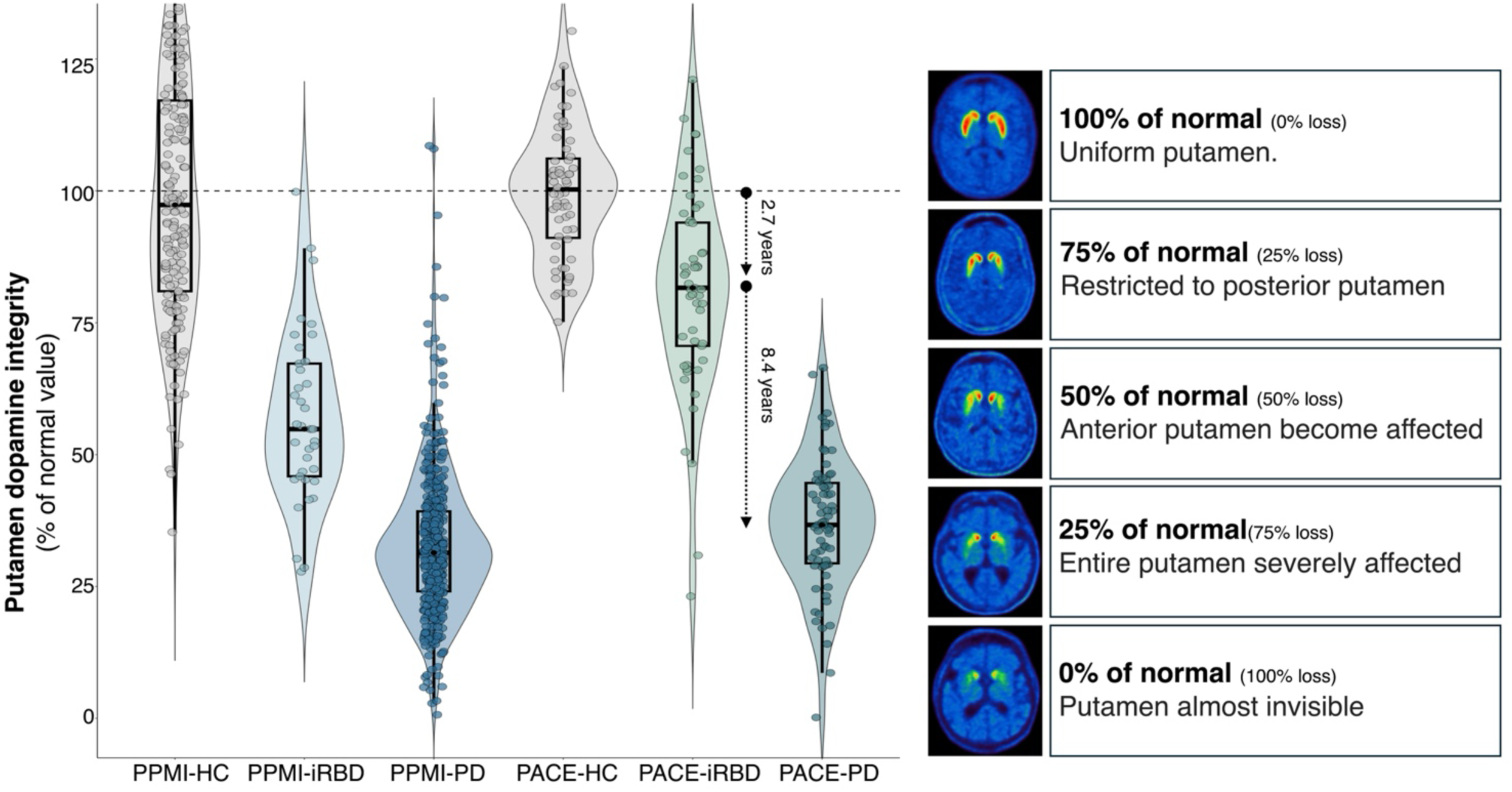
*Left:* Baseline dopaminergic imaging data of most affected putamen presented as % of normal value. The PACE-iRBD cohort show mild dopaminergic degeneration, median = 81.6%, reflecting a preceding degenerative phase of 2.7 years. In comparison, PACE-PD dopaminergic degeneration was 36.7% of normal value. This indicates that the PACE-iRBD cohort will reach the level of PACE-PD in 8.4 years, potentially signifying median time to phenoconversion. One PACE-HC and 23 PPMI-HCs with values high in the normal range are not presented on the figure. *Right:* A representative [^18^F]-FE-PE2I PET image is shown from each degenerative stage - from normal (100%) to almost complete denervation (0%).

**Figure 4.**
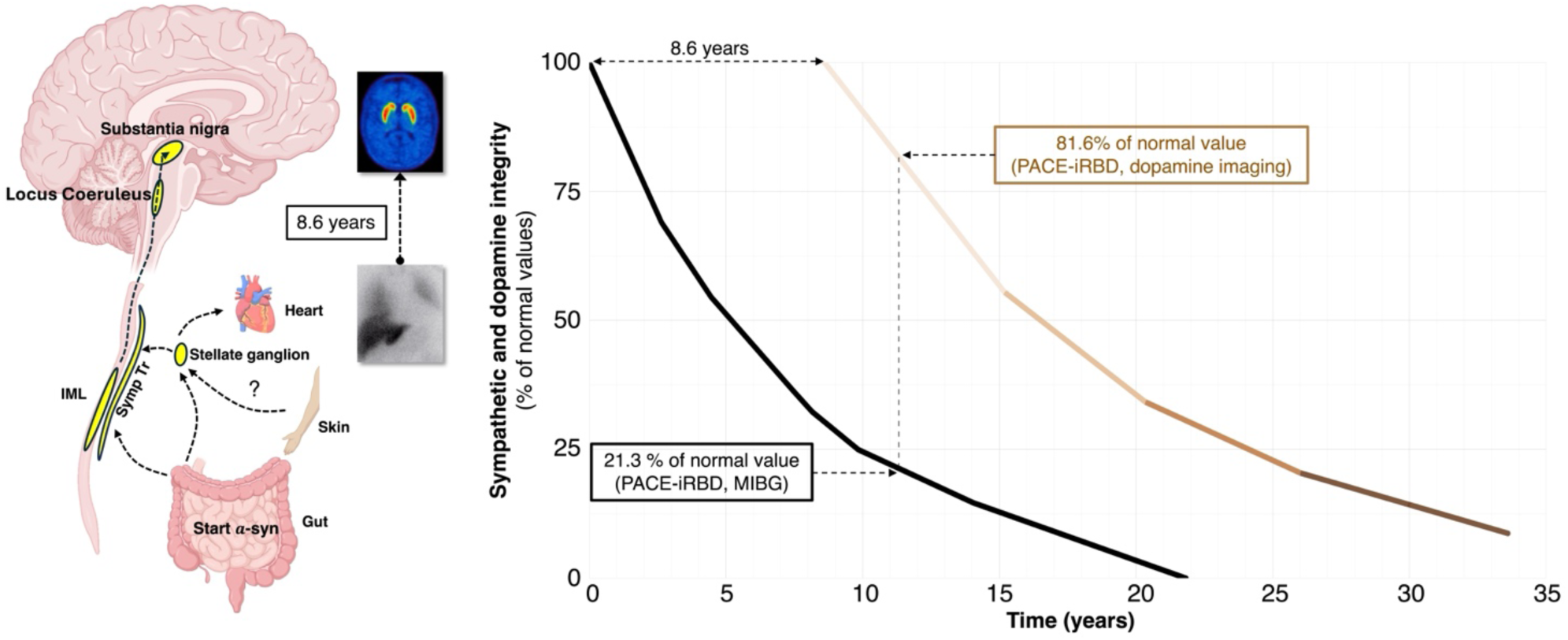
*Left:* Simplified model of proposed α-synuclein propagation through the sympathetic pathways in body-first LBD. Note that the parasympathetic, vagal spreading route has received much more attention in body-first LBD but has been purposefully omitted in this illustration to highlight the importance of sympathetic pathways. *Right*: The cardiac sympathetic (MIBG) and putamen dopamine progression curves with a parallel offset of dopaminergic degeneration based on the baseline PACE-iRBD median MIBG (21.3%) and dopamine imaging (81.6%) results. Notice that onset of dopaminergic degeneration is estimated to begin 8.6 years *later* than cardiac sympathetic degeneration, leaving ample time for a body-first α-synuclein pathology to propagate to brainstem structures and initiate the neurodegenerative process.

### Combined timeline of neurodegenerative events

Based on our findings we can construct an estimated timeline of neurodegenerative events in iRBD (with mean age 67) (**Fig. 5**). At an unknown timepoint before age 56, α-synuclein starts to accumulate and spread within the autonomic nervous system. By median age 56, the cardiac sympathetic system begins to degenerate. Four years later, at age 60, RBD-symptoms begin. By a median age of 64 years, the dopaminergic degeneration starts. The dopaminergic degeneration will reach the level of de novo PD at age 75, predicting the conversion to PD at this age. As the mean age of enrolment in the PACE-iRBD cohort is 67, median time to phenoconversion can be estimated from dopaminergic imaging to 8.4 years. To summarize, the total prodromal period in body-first LBD is therefore estimated to > 19 years. Of note, these estimates are median numbers, which means that some patients experience a shorter prodromal period, whereas others extend to more than 20 years prior to LBD diagnosis.

**Figure 5.**
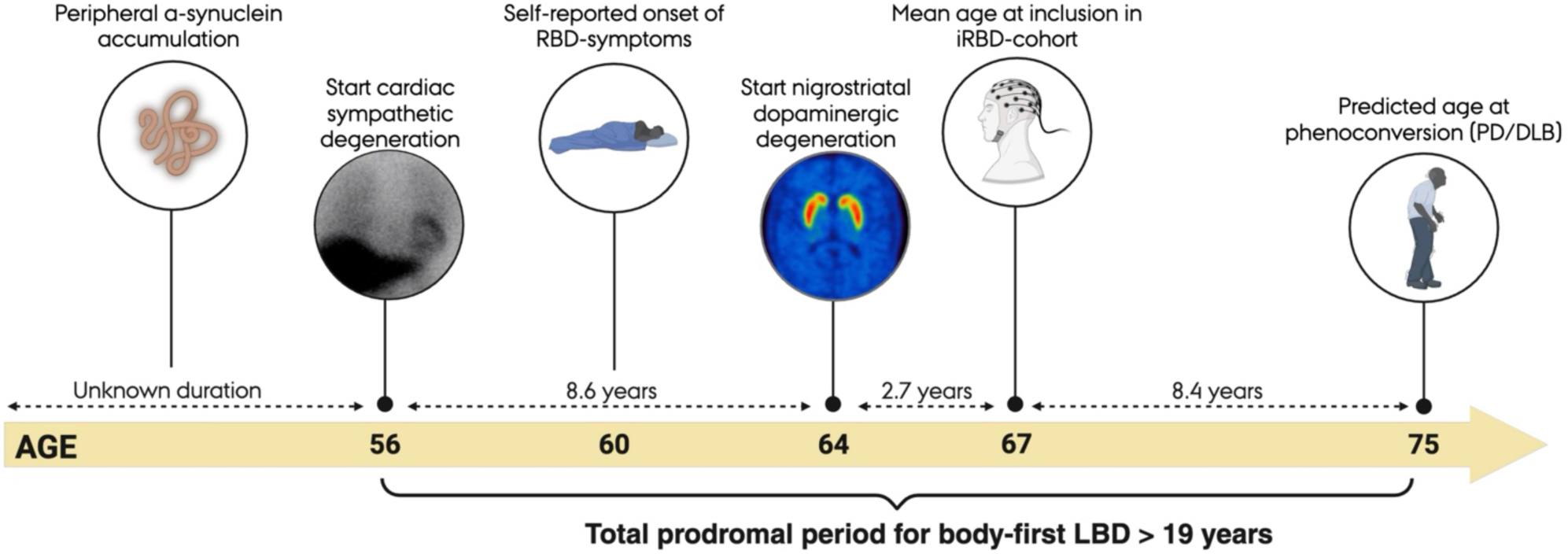
Estimated timeline of neurodegenerative events in body-first LBD, based on progression curves from the present study, and baseline median data from the PACE-iRBD cohort.

## Discussion

In this study, we determined MIBG progression curves from two independent patient cohorts and found nearly identical trajectories (**Fig. 1**). The findings suggest that cardiac denervation shows an initial rapid decline (∼10%-points/year) followed by a slower decline (∼5 %-points/year) towards total denervation. By extrapolating these progression curves, we estimated that cardiac denervation in the PACE-iRBD cohort had started 11.3 years prior to baseline enrolment. The average time to phenoconversion for the PACE-iRBD cohort was estimated to be 8.4 years based on the dopaminergic progression curve. Consequently, the total prodromal period of body-first LBD, where ongoing neurodegeneration can be objectively measured, must in many cases exceed 19 years.

This study is the first to estimate the prodromal period in LBD by modelling the progressive cardiac sympathetic denervation. Yet, the MIBG progression pattern defined here is supported by previous reports. Two studies of more than 200 patients combined found significantly faster annual decline in MIBG uptake in patients with relatively normal baseline uptake compared to those with pathological baseline MIBG^18,19^. Similar results were reported in a study using the cardiac sympathetic PET tracer [^18^F]dopamine, where PD patients without orthostatic hypotension had relatively preserved cardiac sympathetic innervation at baseline but showed rapid progression at 3.8-year follow-up ^20^. In contrast, PD with orthostatic hypotension showed cardiac denervation already at baseline with limited progression at follow-up.

To understand how the onset and progression rate of peripheral sympathetic degeneration in body-first LBD relates to CNS neurodegeneration, we included longitudinal dopaminergic imaging data and applied the same approach. The dopaminergic curve showed similarity to the MIBG curve, but with slightly slower progression (**Fig. 1**), suggesting that cardiac sympathetic neurons exhibit at least similar vulnerability to LBD pathology as nigrostriatal dopaminergic neurons. Dopaminergic degeneration has previously been studied using linear and exponential models based on imaging data, and there are reasons to believe that more flexible, multimodal models are needed to reflect the heterogeneous nature of PD progression ^21,22^. The dopamine trajectory in the present study reproduces previous studies showing a fast initial decline reaching 50% loss in 10-15 years ^23,24^, supporting the robustness of our finding.

Based on the dopaminergic decline rate and baseline dopaminergic difference between PACE-iRBD and PACE-PD patients, the time to phenoconversion was predicted to be 8.4 years. This is in very close agreement with two large longitudinal cohorts, which show 50% phenoconversion after approximately 8 year^2,25^. For the PPMI-iRBD patients, the predicted time to phenoconversion was shorter, estimated to 6.1 years. It is well known that a dopaminergic deficit in iRBD signals incipient phenoconversion^2,26,27^, but here we suggest that the level of dopaminergic degeneration in iRBD patients can predict *when* patients convert. For example, an iRBD patient with a normal dopamine system (∼100% of normal value) is unlikely to convert to PD within the first 10 years. In contrast, a patient with 75% of normal value will most likely convert in 5-10 years, and a patient with 50% of normal value within 5 years (**Fig. 3**). Of note, PD-converters and DLB-converters show similar dopaminergic deficit, indicating that the predictions may be applicable to both groups^26^.

Our study suggests that peripheral α-synuclein pathology initiates more than 11.3 years before study enrolment of many iRBD patients, and hence when these subjects are in their 50’s. This age-group may be an ideal screening population for early body-first LBD detection. Given recent advancement in pathological α-synuclein detection in CSF, skin, and blood using seeding amplification assays (SAA) in iRBD patients^28-31^, these methods may be able to diagnose LBD before overt peripheral neurodegeneration. However, the amount of pathology is probably sparse in the early phase, and it is unknown if current SAA methods are sufficiently sensitive at this stage of disease, although one autopsy study reported positive skin SAA in 6 of 7 incidental LBD cases, i.e., in the preclinical phase^32^. In addition, the Parkinson Associated Risk Syndrome Study found 48% positive cases on CSF SAA in older subjects with a family history of PD and hyposmia^33^.

The recently proposed NSD-ISS system for a biological definition of neuronal α-synuclein disease includes preclinical stages but does not include any markers of peripheral neurodegeneration^34^. Specifically, stage 1A is defined as positive CSF SAA, and stage 1B additionally include dopaminergic deficit (but no clinical signs or symptoms)^34^. Our study suggests that a major lag time between stage 1A and stage 1B exist, estimated here to 8.6 years for body-first LBD subjects (**Fig. 4 and 5**). This aligns with our recent postmortem study, showing that a “sympathetic body-first LBD subtype” accumulates severe levels of α-synuclein pathology in the sympathetic nervous system, before the brainstem is even affected^35^. In this context, cardiac sympathetic imaging methods, such as MIBG and [^18^F]dopamine PET^20,36^, are undoubtedly powerful tools to detect neurodegeneration much earlier than dopamine scans. Indeed, given the present evidence, we propose that an additional stage be added to this framework, i.e., measurable sympathetic neurodegeneration in the absence of dopaminergic degeneration.

These considerations may have important implications for patient selection in future clinical trials of very early SAA positive cases. Such trials should ideally include MIBG (or other markers of peripheral degeneration) to determine which cases eventually develop an LBD, and when this will happen. Also, this approach will limit inclusion of multiple system atrophy cases, who typically show normal cardiac MIBG uptake^8,36,37^, but may be positive on SAA^38^. Furthermore, when neuroprotective treatments become available, iRBD patients would be ideal to receive treatment to postpone or even prevent development of motor and cognitive symptoms. However, iRBD patients still suffer from several non-motor symptoms including symptoms of orthostatic hypotension, urinary dysfunction, erectile dysfunction, constipation and sleep problems^2,7,39,40^ emphasizing the future need for diagnosing body-first LBD even earlier, i.e. before significant injury to the autonomic systems and brainstem has occurred.

This study has several limitations. First, the MIBG progression curves is interpreted in a body-first LBD perspective but was defined from mixed PD cohorts, which had not been stratified into brain-first and body-first subgroups using polysomnography. However, it is currently not possible to determine an MIBG progression curve using only body-first cases, since they often display nearly complete cardiac denervation at study inclusion. Thus, inclusion of brain-first (RBD-negative) cases are necessary to model the upper, more normal part of the MIBG progression curve. Still, a key assumption of our main result is that cardiac denervation follows the same trajectory in both subtypes, which might not necessarily be the case. If the sympathetic degeneration in body-first patients progress faster than brain-first, we may have overestimated the prodromal period in body-first LBD. Second, the two MIBG cohorts did not follow the exact same imaging protocol, as KPD data were obtained 2 hours post injection, whereas PACE MIBG data were obtained 3.5 hours post injection. The impact of this discrepancy is probably limited as one study found nearly identical H/M ratios at 2, 3, and 4 hours postinjection in both LBD subjects and atypical parkinsonism^41^. Nevertheless, converting the values to percentage points relative to healthy controls appears to efficiently mitigate these differences, as the distribution of MIBG data was very similar between the two cohorts (**Fig. 2**). Third, manual delineation of the heart on the 2D scintigraphy may be challenging in the pathological ranges as the heart can be difficult to define and sometimes is completely invisible (**Fig. 2**). This clearly adds noise to the lower part of the curve where the progression is slow or absent, perhaps also explaining why some of these subjects displayed increasing MIBG H/M values at follow-up. Finally, differences in medication between baseline and follow-up may also slightly influence progression rates, although the impact of most common types of medications on MIBG uptake is minimal. Also, subjects on medication with known substantial influence on cardiac MIBG uptake were excluded. However, we cannot rule out that some types of medications have unknown effect on the cardiac MIBG uptake.

In summary, we determined MIBG progression curves, reflecting progressive cardiac sympathetic degeneration, from two separate patient cohorts. Thereby, we estimated that the onset of measurable peripheral neurodegeneration in iRBD patients occurs at least 11.3 years prior to enrolment on average. By adding the estimated time to phenoconversion of median 8.4 years, our study suggests that the total prodromal/preclinical period in body-first LBD exceeds 19 years. Of note, this 19-year timespan comprises the pre-diagnostic stage where active neurodegeneration is ongoing. This is most likely preceded by an unknown time-period where α-synuclein pathology is accumulating but before neurodegeneration ensues. Nevertheless, our study predicts that many body-first LBD patients should become positive on α-synuclein SAA many years before RBD manifests and that MIBG or [^18^F]dopamine cardiac PET are currently the best available tools to detect the earliest signs of neurodegeneration in body-first LBD. This might enable future clinical trials to detect eligible patients earlier, before significant CNS involvement, which could be an ideal situation for current α-synuclein-targeted therapies, since these may be more effective in clearing peripheral aggregates, i.e. outside the blood-brain barrier. In addition, these patients may even be detected before iRBD evolves, mitigating irreversible damage to the peripheral nervous system and brainstem, and delaying the associated autonomic dysfunction and sleep-related problems. Future studies are needed to determine the prodromal period of brain-first, RBD-negative LBD, which we predict is considerably shorter as the dopaminergic system is probably among the first to be affected in this subtype.

## Data Availability

All data produced in the present study are available upon reasonable request to the authors

## Supplementary material

### How we constructed the progression curves

Progression curves were estimated based on subjects with decreasing imaging values at follow-up. Subjects with increasing values were excluded (see discussion in manuscript). Data from each individual subject is represented by a thin line in the supplementary figures (see below). To build the curves, baseline imaging values were divided into quartiles (Q1-Q4), defined as follows:

- Q1: > 75 of normal value
- Q2: 50-75% of normal value
- Q3: 25.50% of normal value
- Q4: 0-25% of normal value

### 1. Start of the curve (Q1)

- The curve begins at the median baseline value of Q1.
- The thick segment of the curve shows the median decline rate in Q1 until either:

I. the curve reaches the last follow-up measurement (on the Time-axis), or
II. the curve reaches the median baseline value of Q2.

### 2. Connecting the segments

Thin lines connect the thick segments by **extrapolation**, using the same decline rate observed in the preceding quartile.

### 3. Q2 segment

- From the median Q2 baseline imaging value, the next thick segment shows the **median decline rate for Q2 subjects.**
- This segment continues until it either reaches the last follow-up measurement (on the Time-axis), or the curve reaches the median baseline value of Q3, i.e., the same process as for Q1.

**Supplementary Figure 1A.**
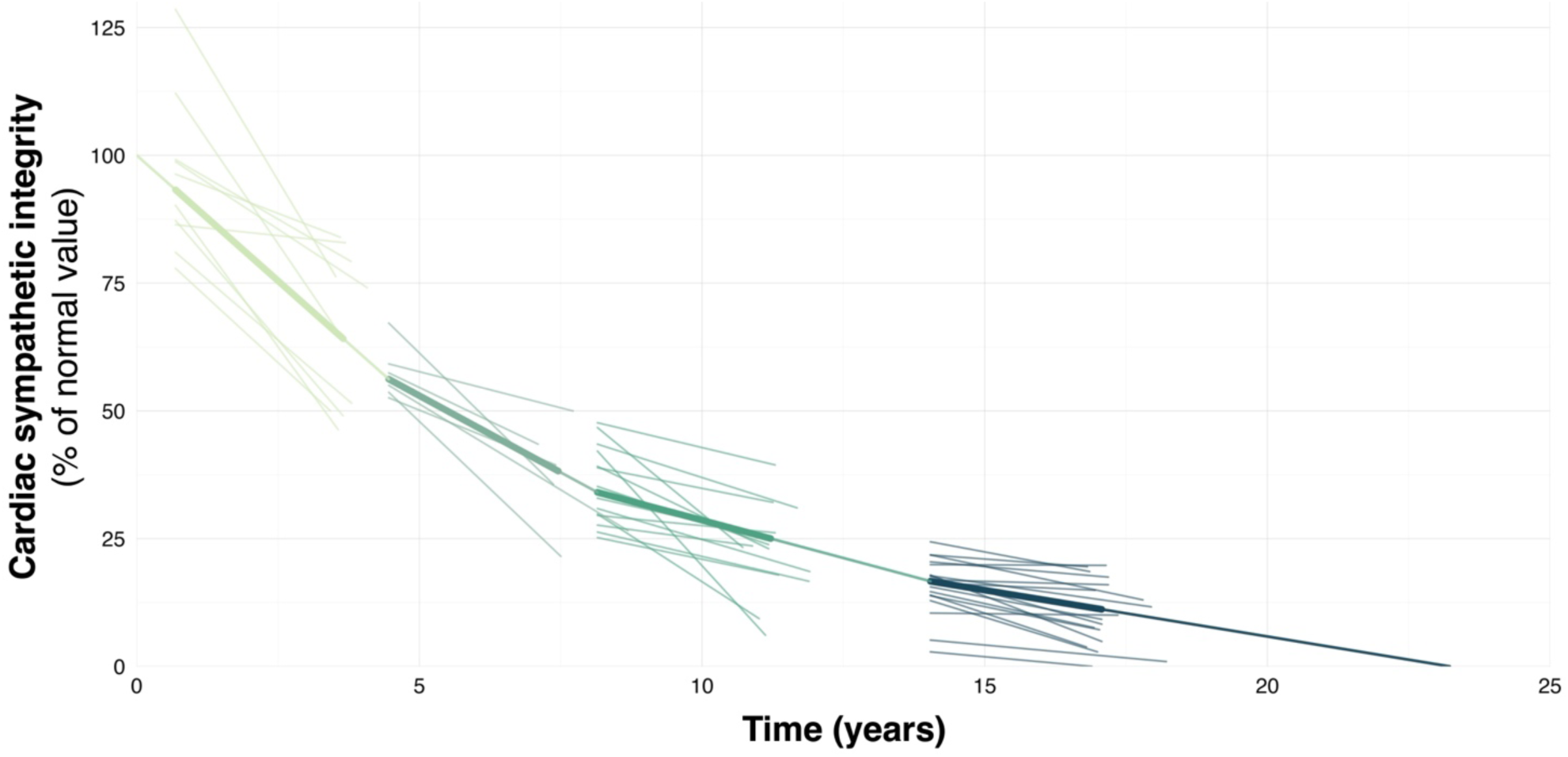
Progression curve and individual MIBG data points from the PACE-cohort.

**Supplementary Figure 1B.**
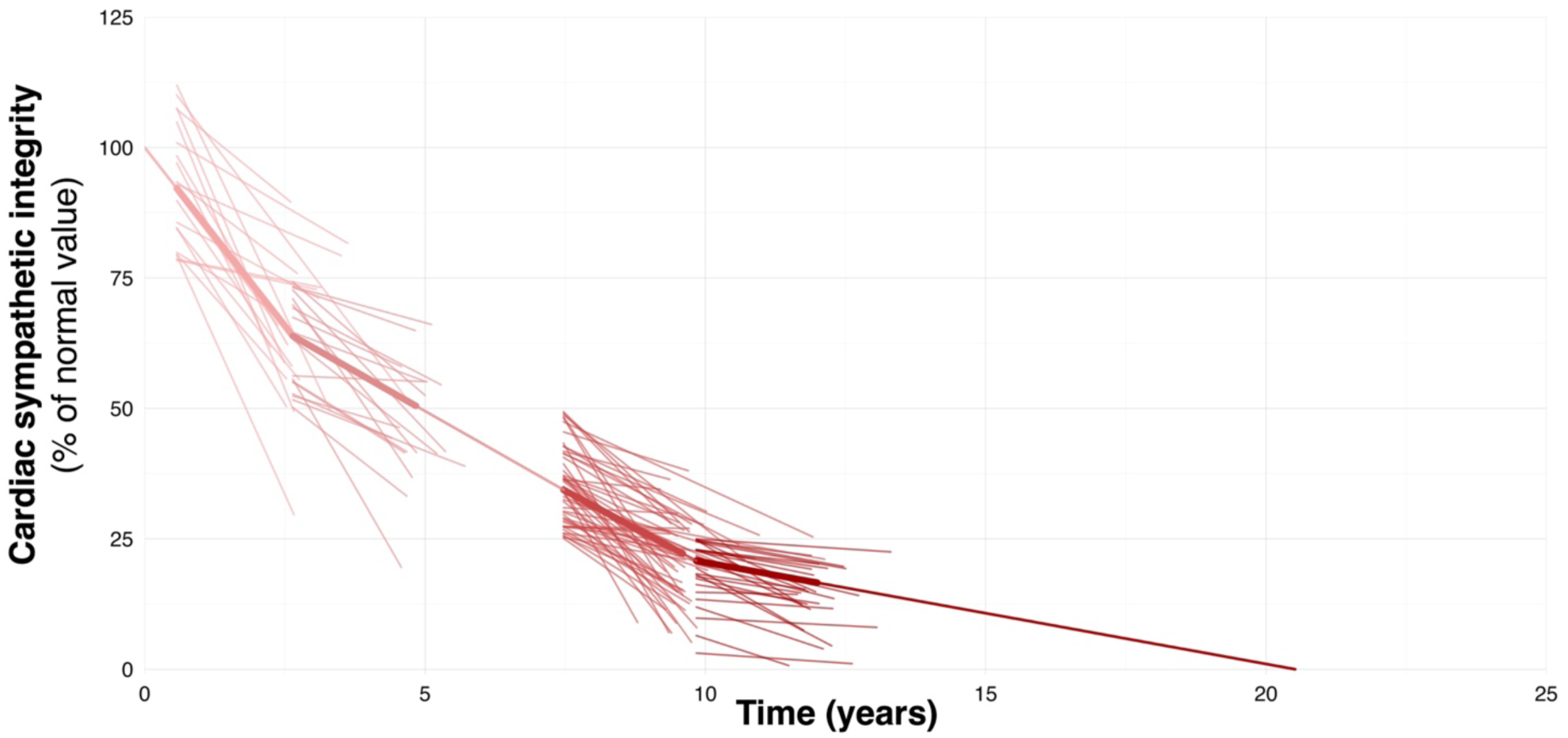
Progression curve and individual MIBG data points from the KPD-cohort. One datapoint with negative H/M Late at follow-up has been omitted from the third quartile on the plot but included in analyses.

**Supplementary Figure 1C.**
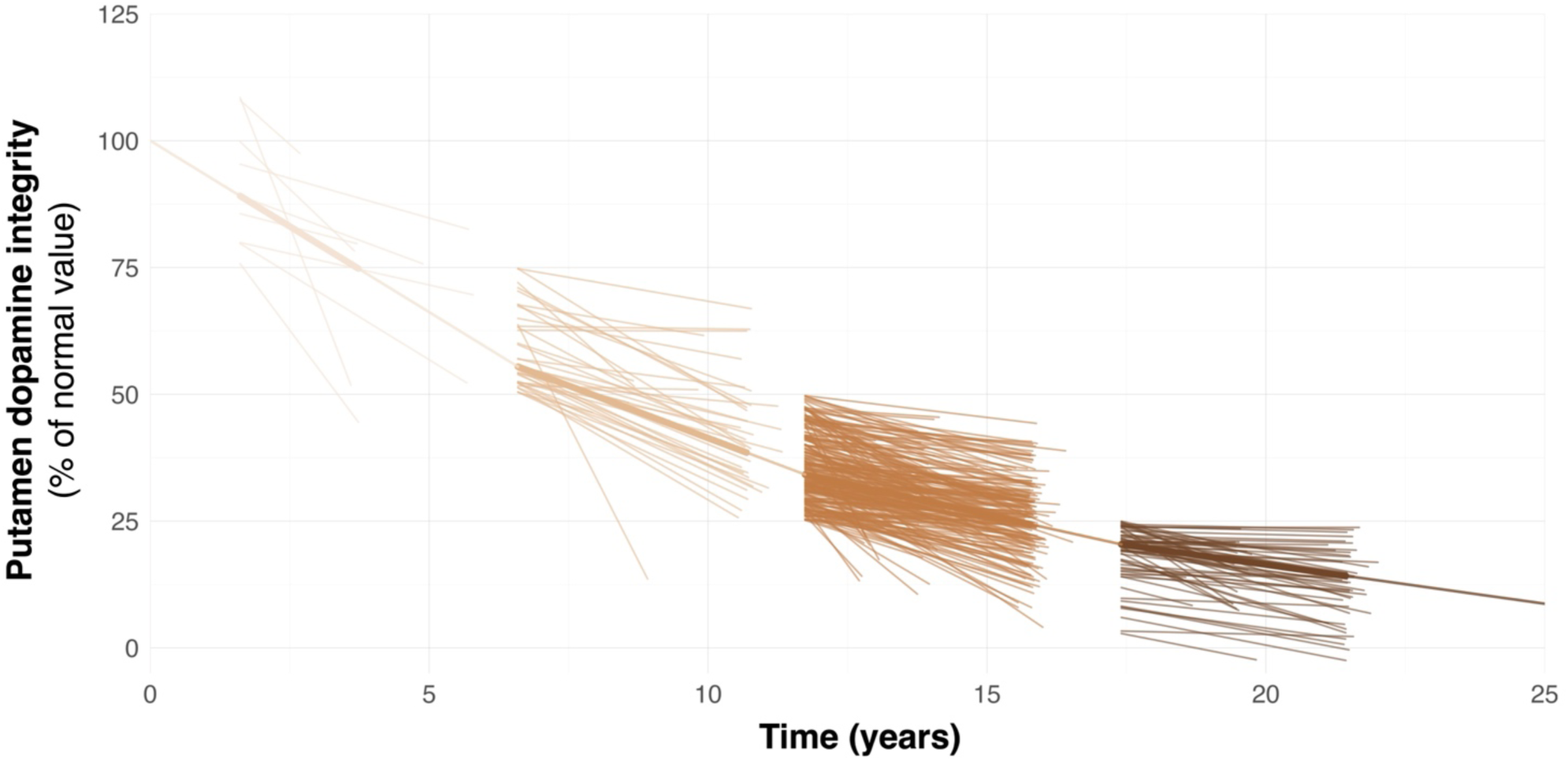
Progression curve and individual DaT data points from the PPMI-cohort.

### 4. Q3 → Q4 → zero

- The same process is repeated for Q3 and Q4 data and extrapolated to reach MIBG = 0%.

The progression curves and individual data points are presented below for PACE MIBG data (**Supplementary Figure 1A**), KPD MIBG data (**Supplementary Figure 1B**), and PPMI DaT data (**Supplementary Figure 1C**).

### Progression curves including subjects with increasing imaging values at follow-up

As discussed in the manuscript, we only included subjects with declining values at follow-up to build the progression curves. In **Supplementary Figure 2A-C** the declining curves including *all* subjects are shown for comparison. Note that all three curves that included subjects with increasing imaging values showed considerable slower progression rates.

**Supplementary Figure 2A.**
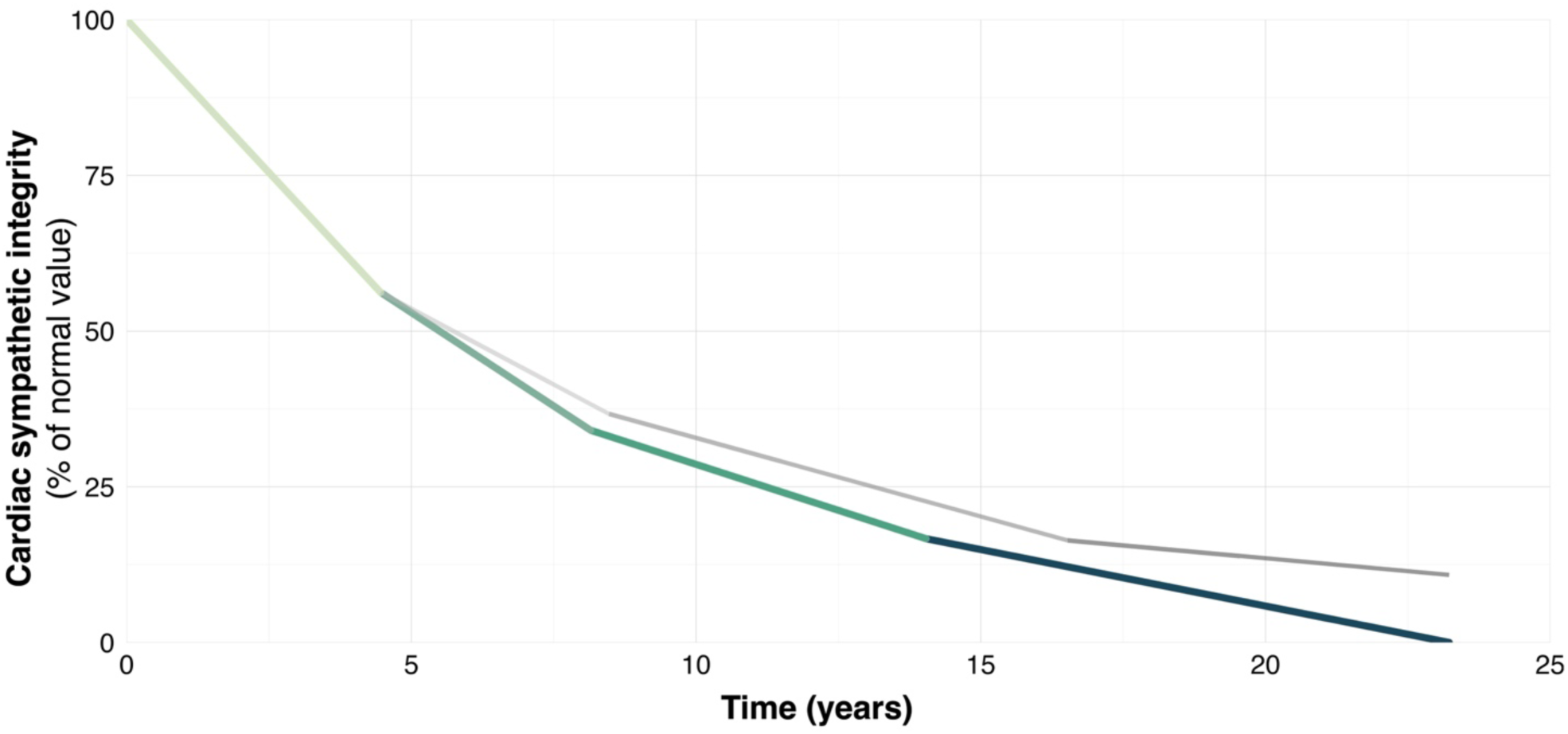
PACE MIBG progression curve with (grey) and without (coloured) subjects with increasing imaging values at follow-up. The coloured curve is identical to Figure 1A (PACE curve) in the manuscript - shown here for comparison.

**Supplementary Figure 2B.**
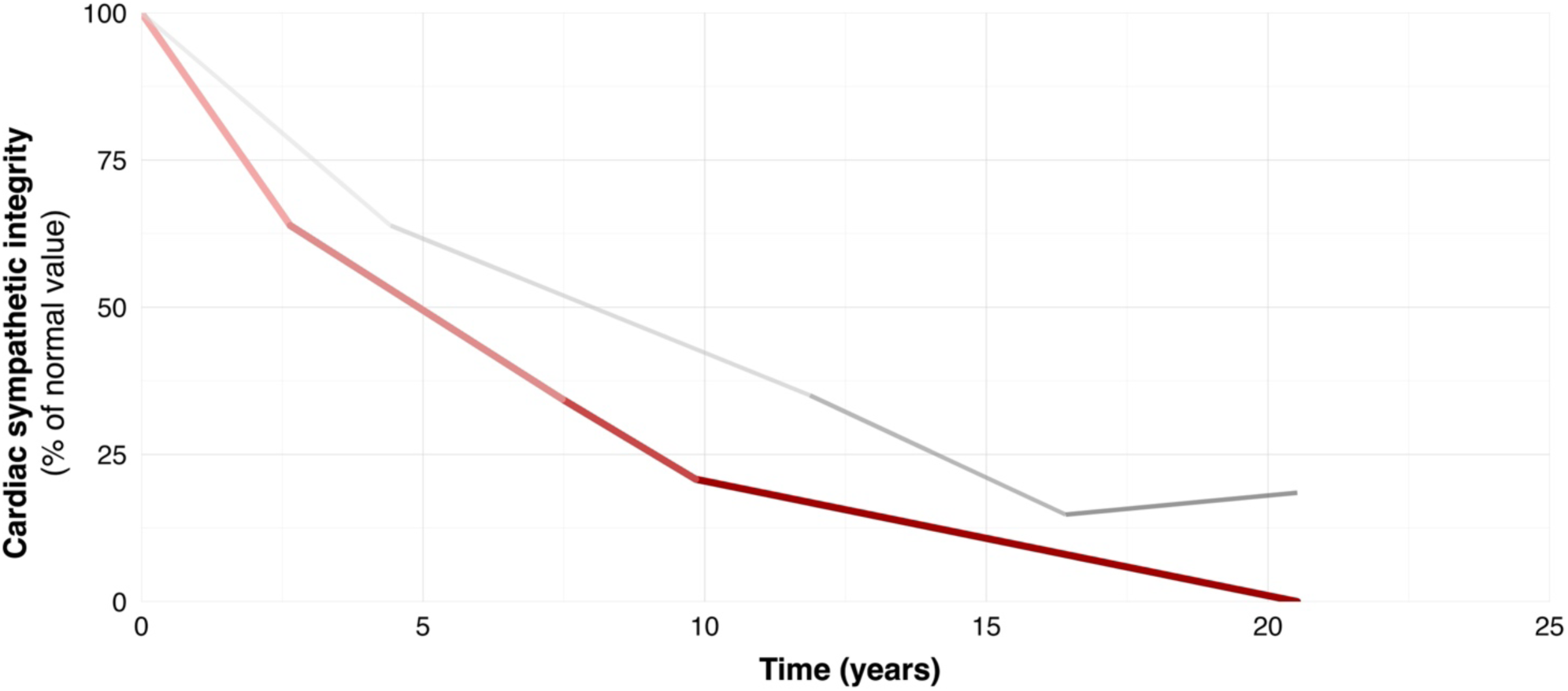
KPD MIBG progression curve with (grey) and without (coloured) subjects with increasing imaging values at follow-up. The coloured curve is identical to Figure 1A (KPD curve) in the manuscript - shown here for comparison.

**Supplementary Figure 2C.**
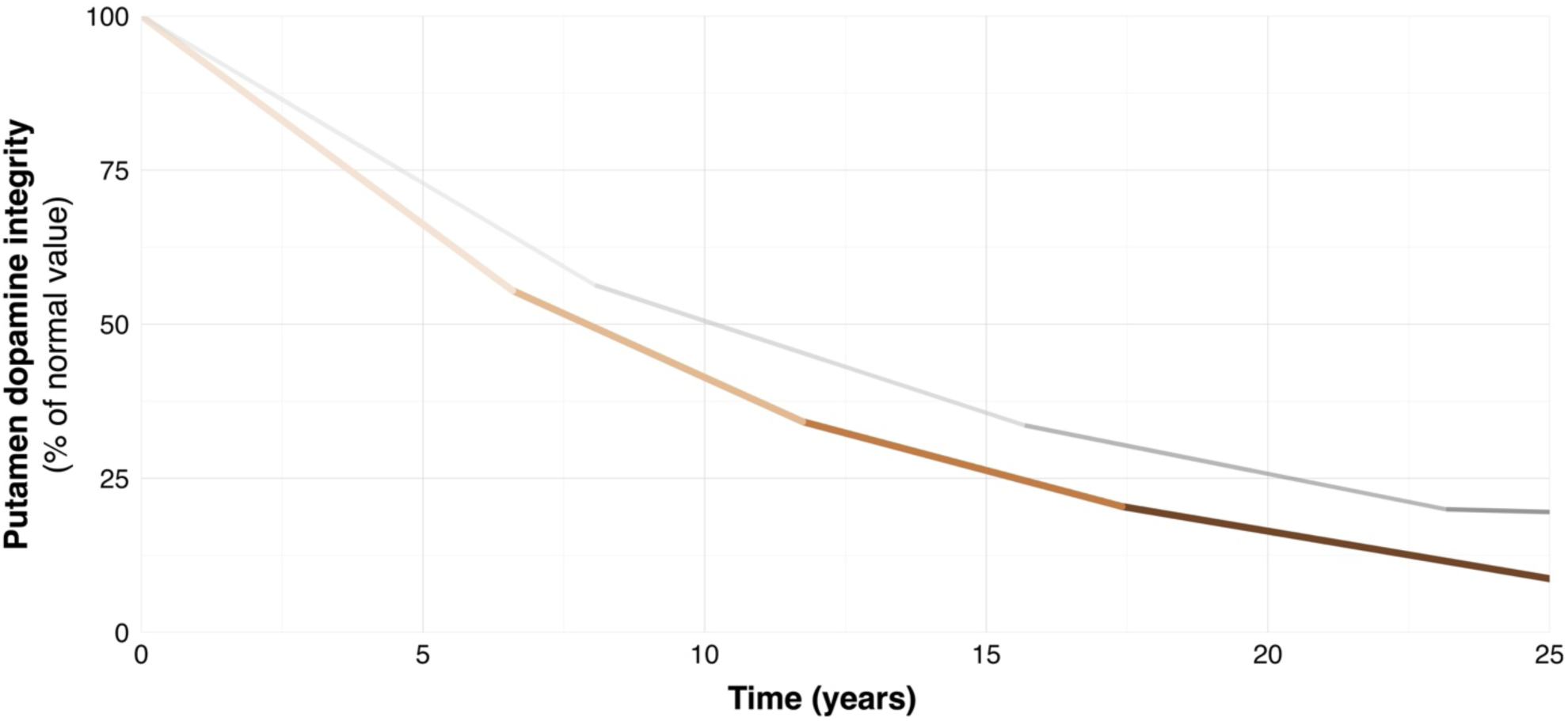
PPMI DaT progression curve with (grey) and without (coloured) subjects with increasing imaging values at follow-up. The coloured curve is identical to Figure 1B in the manuscript - shown here for comparison.

